# A Comparative Effectiveness Study on Opioid Use Disorder Prediction Using Artificial Intelligence and Existing Risk Models

**DOI:** 10.1101/2022.05.18.22275281

**Authors:** Sajjad Fouladvand, Jeffery Talbert, Linda P. Dwoskin, Heather Bush, Amy L. Meadows, Lars E. Peterson, Yash R. Mishra, Steven K. Roggenkamp, Fei Wang, Ramakanth Kavuluru, Jin Chen

## Abstract

**Objective:** To compare the effectiveness of multiple artificial intelligence (AI) models with unweighted Opioid Risk Tool (ORT) in opioid use disorder (OUD) prediction.

**Materials and Methods:** This is a retrospective cohort study of deidentified claims data from 2009 to 2020. The study cohort includes 474,208 patients. Cases are prescription opioid users with at least one diagnosis of OUD or at least one prescription for buprenorphine or methadone. Controls are prescription opioid users with no OUD diagnoses or buprenorphine or methadone prescriptions. Cases and controls are matched based on age, sex, opioid use duration and longitudinal data availability. OUD prediction performance of logistic regression (LR), random forest (RF), XGBoost, long short-term memory (LSTM), transformer, our proposed AI model for OUD prediction (MUPOD), and the unweighted ORT were assessed using accuracy, precision, recall, F1-score and AUC.

**Results:** Data includes 474,208 patients; 269,748 were females with an average age of 56.78 years. On 100 randomly selected test sets including 47,396 patients, MUPOD can predict OUD more efficiently (AUC=0.742±0.021) compared to LR (AUC=0.651±0.025), RF (AUC=0.679±0.026), XGBoost (AUC=0.690±0.027), LSTM (AUC=0.706±0.026), transformer (AUC=0.725±0.024) as well as the unweighted ORT model (AUC=0.559±0.025).

**Discussion:** OUD is a leading cause of death in the United States. AI can be harnessed with available claims data to produce automated OUD prediction tools. We compared the effectiveness of AI models for OUD prediction and showed that AI can predict OUD more effectively than the unweighted ORT tool.

**Conclusion:** Embedding AI algorithms into clinical care may assist clinicians in risk stratification and management of patients receiving opioid therapy.

## INTRODUCTION

Early prediction and engagement of individuals at risk of developing an opioid use disorder (OUD) is a critical unmet need and public health emergency.^1,2^ According to latest estimates, 40.3 million people aged 12 and over in the U.S. have a substance use disorder (SUD), with 2.7 million reporting an OUD.^3^ Individuals with OUD often do not seek treatment or have internalized stigma about OUD that limits treatment.^4,5^ While there are tools developed currently to predict aberrant behavior when prescribing opioids^6^ from a general primary care population^7^, there are only a few clinical tools, such as the Opioid Risk Tool (ORT)^8^ and the revised unweighted version of ORT (ORT-OUD)^9^, developed and validated for assessing the risk of OUD.

However, these tools have been created based on small and unrepresentative samples (ORT using data from 185 patients and ORT-OUD using 1178 patients) and have limited generalizability and predictive validity with sensitivity and specificity ranging from 0.25 to 0.83 and 0.43 to 0.88, respectively.^8,9^ Further, due to the social stigma associated with OUD, primary care professionals are often reluctant to screen and diagnose an individual with OUD using these tools as they require the clinicians to ask about stigmatized events in patients’ history such as personal and family history of substance use and mental health disorders^10^. These complications present barriers for early identification and engagement of individuals at risk for OUD.

Since individuals are reluctant to seek treatment, one solution is to identify patients otherwise engaged in the health care system where a majority of ambulatory care is provided.^11,12^ Available U.S. national administrative and clinical data can be utilized to help clinicians screen and identify high risk patients providing an opportunity for primary care professionals to play a greater role in increasing early detection, treatment, and prevention of OUD. Combining existing and routinely collected administrative data with Artificial Intelligence (AI) offers the potential to understand factors predicting OUD risk and trajectories in a longitudinal framework. To this end, there have been multiple attempts to create OUD prediction tools utilizing healthcare data and AI, machine learning and statistical methods. Classical machine learning and statistical models such as random forests^13^, gradient boosting machines^14^ and logistic regressions^15^ have been used with claims data^16–20^, electronic health records and lab tests^21^ as well as national survey data sets.^22^ Others have produced OUD prediction tools customized for specific cohorts such as lumbar spine surgery patients^23^ or arthroscopic hip surgery patients.^24^ Statewide prescription drug monitoring program data is another source of data that have been used in different parts of the country with higher rates of OUD to create population specific OUD prediction tools.^25,26^ More recently, deep learning models have been applied to solve SUD prediction tasks,^27^ opioid long term use prediction,^28^ as well as OUD prediction^29–32^ and have shown superior performance compared to classical machine learning models.

Although the recently developed AI and machine learning models showed promising performance, they still have significant limitations such as using underrepresented data sets or traditional models with low capacity, and none have been compared with the current risk models for OUD prediction. This study is a comprehensive comparison of the state-of-the-art AI and machine learning models, a novel AI model we have specifically developed to solve OUD prediction tasks, and the ORT-OUD tool as a common practice for OUD risk screening among primary care providers. Our proposed model, called **Mu**lti-stream transformer for **P**redicting **O**pioid use **D**isorder (MUPOD), was developed in our prior pilot work^29^ and is extended in the current study. MUPOD is developed to simultaneously analyze patients’ medications, diagnoses, procedures and demographic records longitudinally by attending to segments within and across these data streams and predicts the onset of OUD.

## METHODS

This retrospective study uses over 11 years of deidentified national claims data to train, test and compare state of the art AI and machine learning models and the ORT-OUD tool as a well-stablished OUD prediction tool. Use of this data for this study was approved as an exempt protocol by The University of Kentucky’s Institutional Review Board.

### Data

The large-scale IBM Health MarketScan Commercial Claims database^33^ (formerly known as Truven) including years 2009 to 2020 were used. Data include person-specific clinical utilization, expenditures and enrollment across inpatient, outpatient, prescription drug and carve-out services. The database contains over 3 billion prescriptions, 8 billion diagnoses, and 10 billion procedure records for over 164 million patients. These enrollees are nationally representative of the US population with respect to gender, regional distribution, and age.

### Cohort Selection

The study cohort for this work includes patients with 1) at least one OUD diagnosis (ICD-9 of 304.0x, 305.5x or ICD-10 of F11.xx) in their records or at least one prescription for medications used to treat opioid use disorder, buprenorphine or methadone, and 2) at least 3 opioid prescriptions (other than buprenorphine and methadone) filled six months or more prior to the first OUD diagnoses date, and 3) at least 12 months of data availability. The inclusion criteria of at least 3 opioid prescriptions and 12 months of data availability were designed based on domain knowledge and to address data sparsity and outlier issues in the data. This selection resulted in 237,104 cases. The control cohort includes patients who: 1) have never been diagnosed with OUD nor received a buprenorphine or methadone prescription, and 2) have at least 3 opioid prescriptions (other than buprenorphine and methadone) filled six months or more prior to their last record in the data, 3) have at least 12 month of data availability. This selection resulted in 7,755,649 controls. Cases and controls were then matched based on age, sex, opioid use duration and longitudinal data availability using an anchor-based method (refer to section S2 in the supplement for more details on our matching algorithms). The final data includes 474,208 patients with equal number of cases and controls which were used to conduct our experiments.

Our experiments include both balanced and imbalanced scenarios (with substantially more controls than cases to assess real world utility of our methods). Note, the prediction window in this study is 6 months meaning that the prediction is performed using data from the first record up to 6 months prior to initial diagnosis/last record date.

### Predictors

Predictors included all medications, diagnoses and procedure codes as well as demographic variables including sex and age. Medications were grouped using their root classification based on Medispan generic product identifiers^34^, and diagnoses and procedures were grouped using clinical classification software (CCS)^35^ codes. Medications, diagnoses and procedures included 94, 283 and 242 variables, respectively plus the 2 demographic variables. Variable distributions were used to exclude extremely sparse variables (see section S3 in supplemental material). The final variable set contained 269 variables including 50 medications, 138 diagnoses, 79 procedures, and 2 demographic variables.

### AI models

Machine learning models used in this study included logistic regression^15^, random forest^13^ and xgboost^14^ methods. Deep learning models include long short-term memory^36^ model, transformer model^37^ and an extended version of the novel deep learning model we developed for OUD prediction called MUPOD.^29^ MUPOD was originally developed for OUD prediction using 22 medications and diagnoses codes for 392,492 patients with spondylosis; intervertebral disc disorders; other back problems (CCS code of 205). Here, we extended this method and trained it using 269 medications, diagnoses, procedures and demographics of 474,208 patients’ data.

Section S5 in the supplemental material includes more details on the MUPOD’s architecture and mathematical equations.

LR and RF were trained using static data as they are more effective on static and non-longitudinal data. LSTM, Transformer and MUPOD were trained using longitudinal data (see section S1 in the supplemental materials for more details on data formatting). The longitudinal data for each patient *p*_*i*_ in a complete list of *n* patients *P* = {*p*_1_, …, *p*_*n*_} includes tuples 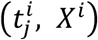 Where 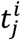 is the *j*^th^ timestamp for patient *p*_*i*_ And 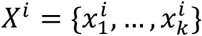 is a vector including medications, diagnoses, procedures and demographics information of patient *p*_*i*_ at time 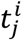. To create static date, this format was converted to a matrix Y with *P* = {*p*_1_, …, *p*_*n*_} rows and *F* ={*f*_1_, …, *f*_*n*_} columns where *P* is the number of patients, and *F* is the number of static variables including frequency values for medication, diagnosis, procedures features across time steps as well as the demographic features. LSTM models were trained using concatenated medication, diagnoses, procedure and demographic data streams. We dynamically unrolled the LSTM models based on the input sequences’ lengths and applied a fully connected layer and an argmax function on the last output to make the final decisions^38^. Transformer is the original encoder block of the transformer model^37^ trained using concatenated medication, diagnosis, procedures and demographics features. Then, a fully connected layer and softmax^39^ function were used to perform the final classifications.

### Performance assessment

Accuracy, precision, recall, F1-score and area under the receiver operating curve (AUC) were used to measure the predictive power of all models. Accuracy measures the ratio of total correct predictions of the models for all samples in the test sets. Precision (also known as positive predictive value) shows how precise a model is when predicting a sample as OUD positive. Recall (or sensitivity) shows how accurate the model is in retrieving all OUD positive samples in the test data. F1 is the harmonic mean of precision and recall and AUC summarizes the model’s predictive modeling capabilities across different settings.^40^ All models were tested on 100 randomly selected test sets sampled from 47,396 unseen patients’ data. The results presented in this study are averages and standard deviations across these 100 test sets.

## RESULTS

### Sample characteristics

In general, demographics in terms of age, sex, months of opioid use prescription, and data availability were similar among cases and controls (Table 1). For example, average age of cases and controls were 56.51 years (25^th^ and 75^th^ percentiles = 46, 66) and 56.95 years (25^th^ and 75^th^ percentiles = 46,66), respectively. The majority were female (56.89% among both cases and controls), cases had a higher average of opioid prescription use (93.54 MME/day among cases vs. 73.22 MME/day among controls).

**Table 1.**
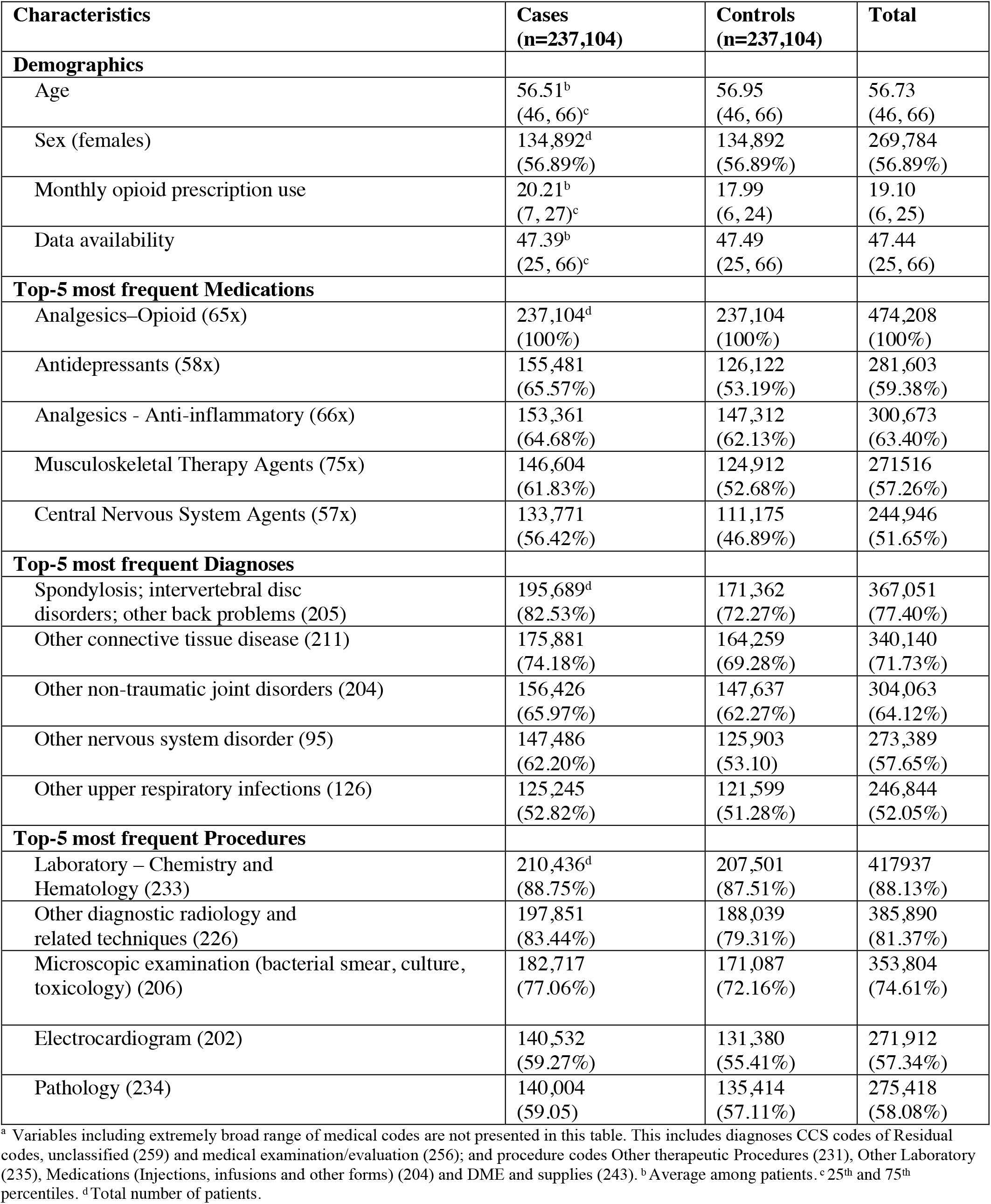
Patient Characteristics among Cases and Controls, IBM MarketScan, 2009 – 2020. Demographic variables, and top-5^a^ frequent medications, diagnoses and procedures among cases.

The biggest difference between the top-5 medication use frequencies among cases and controls included antidepressant medications (65.57% vs 53.19%, respectively). In terms of top-5 most frequent diagnoses, the “Spondylosis; intervertebral disc disorders; other back problems (CCS code = 205)” was more common among cases compared to controls (82.53% vs 72.27%). For procedures, “Microscopic examination (bacterial smear, culture, toxicology) (CCS code = 206)” was more common among cases than controls (77.06% vs 72.16%).

### OUD prediction on balanced test sets

Prediction results for logistic regression, random forest, XGBoost, LSTM, transformer and MUPOD as well as the ORT-OUD^9^ tool are presented in Table 2. Note, the suggested decision threshold for ORT-OUD is 3; however, the prediction performance results for thresholds 1, 2, 3 and 4 provide a more comprehensive comparison. The models corresponding to each of these thresholds are denoted as ORT-OUD-x where x=1, 2,3, or 4. MUPOD has the highest accuracy (0.652±0.019), recall (0.890±0.021), F1-score (F1=0.718±0.014) and AUC (0.742±0.021) compared to all other AI models and the ORT-OUD tools. Ranking the models in Table 2 from the most efficient to the least with regard to their AUC scores as a primary metric for model evaluation reveals MUPOD > transformer > LSTM > XGBoost > random forest> logistic regression> ORT-OUD. Among all versions of ORT-OUD tools, the highest performing threshold turned out to be 1 instead of 3 in terms of F1-score. As the thresholds increases from 1 to 4, the precision increases while the recall significantly drops leading to a significantly lower F1 score.

**Table 2.** Performance of OUD prediction using AI, machine learning and ORT-OUD models.

| Model | Acc. | Prec. | Rec. | F1 | AUC |
| --- | --- | --- | --- | --- | --- |
| Logistic Regression | 0.609<br>±0.021 | 0.631<br>±0.026 | 0.519<br>±0.031 | 0.569<br>±0.027 | 0.651<br>±0.025 |
| Random Forest | 0.631<br>±0.022 | 0.624<br>±0.022 | 0.650<br>±0.030 | 0.637<br>±0.022 | 0.679<br>±0.026 |
| XGBoost | 0.638<br>±0.023 | 0.631<br>±0.024 | 0.659<br>±0.031 | 0.644<br>±0.024 | 0.690<br>±0.027 |
| LSTM | 0.648<br>±0.024 | 0.628<br>±0.022 | 0.719<br>±0.033 | 0.670<br>±0.023 | 0.706<br>±0.026 |
| Transformer | 0.651<br>±0.021 | 0.603<br>±0.016 | 0.874<br>±0.023 | 0.713<br>±0.016 | 0.725<br>±0.024 |
| MUPOD | **0.652<sup>a</sup>**<br>±0.019 | 0.602<br>±0.014 | **0.890**<br>±0.021 | **0.718**<br>±0.014 | **0.742**<br>±0.021 |
| ORT-OUD-1 | 0.534<br>±0.021 | 0.523<br>±0.015 | 0.714<br>±0.030 | 0.603<br>±0.019 | 0.559±<br>0.025 |
| ORT-OUD-2 | 0.547<br>±0.020 | 0.574<br>±0.034 | 0.350<br>±0.030 | 0.434<br>±0.031 | 0.559±<br>0.025 |
| ORT-OUD-3<sup>b</sup> | 0.527<br>±0.013 | 0.619<br>±0.064 | 0.129<br>±0.020 | 0.213<br>±0.030 | 0.559±<br>0.025 |
| ORT-OUD-4 | 0.513<br>±0.007 | **0.691**<br>±0.131 | 0.039<br>±0.011 | 0.074<br>±0.021 | 0.559±<br>0.025 |

<sup>a</sup> Bolded numbers are the highest values in each column. <sup>b</sup> Proposed decision threshold in the original ORT-OUD study is 3.

### OUD prediction on imbalanced test sets

The predictive power of the models was further tested on three imbalanced test sets to estimate the generalizability of the models more efficiently in real world environment (Table 3). We created three imbalanced test sets with the ratio of cases to controls set to 0.5, 0.2, 0.1 and 0.01 and tested the models using these imbalanced test sets. The highest AUC for all imbalanced test sets was obtained by MUPOD. The ORT-4 had the highest precisions (0.517±0.209, 0.304±0.204, 0.156±0.192 and 0.020±0.064 for .5N, .2N, .1N and .01N test sets, respectively) and the MUPOD model had the highest recall (0.888±.029, 0.883±.047, 0.897±.058, and 0.918±0.180 for .5N, .2N., .1N and .01N test sets, respectively). MUPOD had the highest F1 score (0.579±.018) for the .5N test sets, but the highest F1 score for the other three imbalanced test sets were obtained using the LSTM model (0.370±0.031, 0.245±0.030 and 0.035±0.014 for .2N, .1N, and .01N, respectively).

**Table 3.** OUD prediction results for imbalanced test sets.

| Model | Precision |  |  |  | Recall |  |  |  | F1-Score |  |  |  | AUC |  |  |  |
| --- | --- | --- | --- | --- | --- | --- | --- | --- | --- | --- | --- | --- | --- | --- | --- | --- |
|  | .5N<sup>a</sup> | .2N | .1N | .01N | .5N | .2N | .1N | .01N | .5N | .2N | .1N | .01 | .5N | .2N | .1N | .01 |
| Logistic Regression | .461<br>±.036 | .254<br>±.032 | .147<br>±.025 | .016<br>±.010 | .518<br>±.044 | .517<br>±.076 | .522<br>±.100 | .478<br>±.315 | .487<br>±.035 | .340<br>±.044 | .228<br>±.039 | .030<br>±.020 | .650<br>±.030 | .652<br>±.046 | .656<br>±.052 | .646<br>±.167 |
| Random Forest | .454<br>±.029 | .248<br>±.024 | .145<br>±.022 | .016<br>±.008 | .651<br>±.045 | .644<br>±.064 | .663<br>±.102 | .652<br>±.295 | .535<br>±.031 | .358<br>±.034 | .238<br>±.035 | .032<br>±.016 | .679<br>±.030 | .675<br>±.039 | .686<br>±.057 | .676<br>±.162 |
| XGBoost | .461<br>±.030 | .252<br>±.025 | .148<br>±.020 | .018<br>±.009 | .657<br>±.042 | .650<br>±.067 | .668<br>±.096 | .690<br>±.292 | .541<br>±.030 | .363<br>±.035 | .242<br>±.032 | .034<br>±.017 | .689<br>±.031 | .689<br>±.038 | .699<br>±.051 | .693<br>±.162 |
| LSTM | .458<br>±.024 | .251<br>±.021 | .147<br>±.018 | .018<br>±.007 | .718<br>±.043 | .712<br>±.067 | .733<br>±.091 | .775<br>±.292 | .559<br>±.027 | .370<br>±.031 | .245<br>±.030 | .035<br>±.014 | .705<br>±.029 | .705<br>±.042 | .717<br>±.057 | .724<br>±.167 |
| Transformer | .431<br>±.018 | .233<br>±.015 | .132<br>±.012 | .015<br>±.005 | .872<br>±.031 | .871<br>±.048 | .872<br>±.071 | .855<br>±.250 | .577<br>±.020 | .367<br>±.022 | .229<br>±.020 | .029<br>±.011 | .724<br>±.026 | .721<br>±.036 | .724<br>±.052 | .720<br>±.156 |
| MUPOD | .430<br>±.016 | .231<br>±.012 | .132<br>±.009 | .015<br>±.004 | **.888<sup>b</sup>**<br>±.029 | **.883**<br>±.047 | **.897**<br>±.058 | **.918**<br>±.180 | **.579**<br>±.018 | .366<br>±.018 | .230<br>±.016 | .030<br>±.009 | **.741**<br>±.023 | **.737**<br>±.035 | **.746**<br>±.052 | **.732**<br>±.136 |
| ORT-ODD-1 | .354<br>±.016 | .180<br>±.016 | .099<br>±.012 | .011<br>±.005 | .713<br>±.035 | .716<br>±.069 | .712<br>±.086 | .730<br>±.303 | .472<br>±.020 | .288<br>±.026 | .173<br>±.021 | .022<br>±.009 | .556<br>±.027 | .561<br>±.042 | .560<br>±.058 | .562<br>±.188 |
| ORT-ODD-2 | .399<br>±.035 | .213<br>±.036 | .121<br>±.031 | .013<br>±.011 | .344<br>±.039 | .351<br>±.063 | .357<br>±.095 | .337<br>±.305 | .369<br>±.034 | .265<br>±.044 | .180<br>±.046 | .024<br>±.022 | .556<br>±.027 | .561<br>±.042 | .560<br>±.058 | .562<br>±.188 |
| ORT-ODD-3* | .440<br>±.080 | .255<br>±.084 | .143<br>±.073 | .018<br>±.030 | .125<br>±.029 | .135<br>±.048 | .133<br>±.073 | .128<br>±.208 | .194<br>±.042 | .175<br>±.059 | .136<br>±.071 | .031<br>±.051 | .556<br>±.027 | .561<br>±.042 | .560<br>±.058 | .562<br>±.188 |
| ORT-ODD-4 | **.517**<br>±.209 | **.304**<br>±.204 | **.156**<br>±.192 | **.020**<br>±.064 | .037<br>±.018 | .039<br>±.028 | .036<br>±.042 | .040<br>±.123 | .069<br>±.032 | .068<br>±.048 | .057<br>±.065 | .026<br>±.082 | .556<br>±.027 | .561<br>±.042 | .560<br>±.058 | .562<br>±.188 |

<sup>a</sup> The .xN means the number of samples in the cases are 0.x times smaller than the number of samples in controls. <sup>b</sup> Bolded numbers are the highest values in each column. \* The recommended decision threshold for ORT-ODD model is 3.

Table 3 also shows the effect of using different decision thresholds for the ORT-OUD tool. Among the ORT-OUD tools, the ORT-OUD-1 had the highest F1 score for two of the imbalanced test sets (0.472±0.020 and 0.288±0.026 for .5N and .2N sets, respectively). ORT-OUD-3, the ORT-OUD tool with the recommended threshold of 3, had the highest F1 score (0.180 ± 0.046) for the most extreme imbalanced test set. However, ORT-OUD-4 would be a more effective tool compared to the other versions of ORT-OUD tools when a more precise decision-making system is desired.

Overall, MUPOD with the highest AUC for all balanced and imbalanced test sets had the best performance compared to all other models. ORT-OUD-4 was the most precise model, but with the cost of significantly sacrificing recall and F1scores. Note, similar to ORT-OUD models, all AI models can be configured in favor of producing very high precision using the decision-making threshold in their final neural layer. Precision at recall 0.50 for MUPOD on the 0.5N, 0.2N, 0.1N, and 0.01N sets are 0.557±0.036, 0.326±0.043, 0.205±0.044, and 0.022±0.015, respectively, which are significantly higher than relevant OUD-ORT-4 while recall 0.500 is still higher than the ORT-OUD-4 recalls too. Therefore, MUPO can be set in favor of precision if needed and still performs significantly more effective than all the ORT tools in Table 3.

### Effect of morphine milligram equivalent

The effect of opioid dosage prescribed to those patients in the test sets that MUPOD could successfully classify as cases (true positive) and controls (true negative) was explored. Dosage was measured using Morphine Milligram Equivalent (MME) per day^41^. Table 4 presents MME statistics for true positive and true negative cohorts. Setting A in Table 4 presents the total average MME per day across all records from the first record to the first diagnosis date or last record date. Further, average MME is shown for the records within the 6-month prediction window prior to the first OUD diagnosis/last record date. The total average MME per day for cases was 93.54 (30.72, 25^th^ percentile and 68.45, 75^th^ percentile), while this number for controls was 73.22 (24.67, 25^th^ percentile and 48.91, 75^th^ percentile). This difference is significantly greater the closer to OUD onset (within the 6-month prediction window). For cases, this number was 94.05 (20.32, 25^th^ percentile and 67.50, 75^th^ percentile) versus 55.19 (0, 25^th^ percentile and 39, 75^th^ percentile) for controls. A similar gap between opioid dosage prescription for cases and controls has been reported repeatedly in the literature.^42–46^ In setting B, accumulated value for all opioid prescription for each patient was computed and averaged across all patients in each cohort. Sum of MME prescribed to cases was 5452.07 while this number for controls was 2906.61 and this difference is significantly greater within the 6-month prediction window; 984.13 for cases versus 433.16 for controls.

**Table 4.** Morphine Milligram Equivalent (MME) per day computed for the true positive and true negative cohorts.

| Characteristics | True Positives | True Negatives |
| --- | --- | --- |
| n | 20881 | 9886 |
| Age | 56.52 (46, 66) | 57.04 (47, 66) |
| Female | 13,337 (56.57%) | 13,624 (57.20%) |
| **Setting A** (average MME) |  |  |
| Averaged MME: all records (from first record to OUD diagnoses/last record date) | 93.54 (30.72, 68.45) | 73.22 (24.67, 48.91) |
| Averaged MME: within the 6-month prediction window immediately preceding the diagnoses/last record | 94.05 (20.32, 67.50) | 55.19 (0.00, 39.00) |
| **Setting B** (accumulated MME) |  |  |
| Accumulated MME: all records (from first record to OUD diagnoses/last record) | 5452.07 | 2906.61 |
| Accumulated MME: within the 6-month prediction window immediately preceding the diagnoses/last record | 984.13 | 433.16 |

The numbers for MME are overall average (25^th^ percentile, 75^th^ percentile).

### Risk factor identification

We used the shapely additive explanation (SHAP)^47^ to perform feature importance assessments using XGBoost model (see section S6 for more details on the XGBoost model’s setting) (Figure 1). SHAP works best with our XGBoost model as our current version of MUPOD is not compatible with SHAP. Blue indicates lower frequencies and red indicates higher frequencies of the variables; negative SHAP values show association with lower risks of OUD and positive values show association with higher risks of developing OUD. The fewer the diagnoses and procedures such as ‘spondylosis; intervertebral disc disorders, other back problems’, ‘other diagnostic procedures (interview, evaluation, consultation)’, the lower the risk of developing OUD; likewise, the fewer the prescriptions for ‘neuromuscular agents’, ‘antidepressants’, ‘central nervous system agents’ and ‘analgesics-opioid’ the smaller the risk for OUD. Increasing age, presence of ‘other screening for suspected conditions’ and ‘essential hypertension’ diagnoses were negatively associated with OUD. These findings is consistent with past related studies.^48–50^

**Figure 1.**
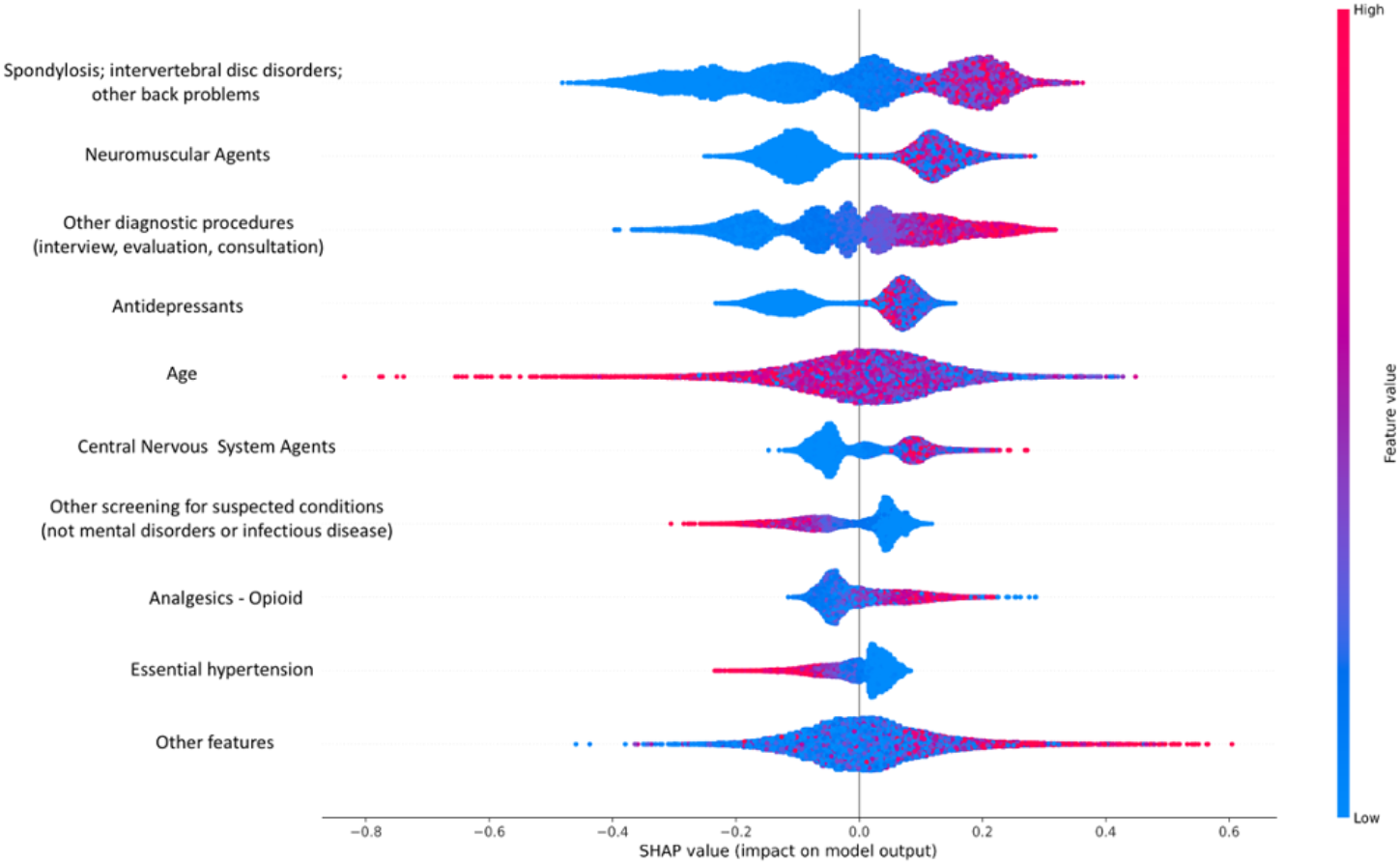
OUD risk factor identification using shapely additive explanation method. Blue indicates lower frequencies and red indicates higher frequencies of the variables; negative SHAP values show association with lower risks of OUD and positive values show association with higher risks of developing OUD.

## DISCUSSION

Using the large-scale national IBM MarketScan data to train and test AI models and compare their performance with the ORT-OUD tool, we found that AI models better predicted OUD 6 months before the onset of the disease. The proposed AI model, MUPOD, had a higher AUC and F1-score compared to the ORT-OUD tool on the balanced test set. Further, MUPOD had a higher AUC and F1 score on all imbalanced test sets compared to the ORT-OUD tools.

Our findings suggest that using an AI algorithm to identify OUD in clinical care may be more effective, and less intrusive, than standard clinical pathways.

At present, healthcare providers use ORT based tools with the patient in front of them, asking questions interactively about risk for OUD. There are limitations with this current state including willingness of patients to answer sensitive questions due to stigma associated with OUD and with personal and family history. Our study demonstrates that all of the items in ORT-OUD tool can be efficiently extracted from the large-scale data with automatic extraction of pertinent medical history. Therefore, we believe using claims data to assess patients’ risk of developing OUD is still a promising heuristic for using the ORT-OUD tool and could potentially be combined with individual patient encounters and information to inform clinical diagnosis.

## LIMITATION

Although AI models were thoroughly evaluated using repeated testing, more studies are needed to assess the current AI model further in two ways. First, testing the current model using claims data sets other than IBM MarketScan. Second, more prospective studies are needed to assess our AI model in a clinical setting where the AI model and the ORT based tools are used to screen the OUD risk and the patients are followed up for OUD diagnoses longitudinally. In this study, we used prescriptions, diagnoses, procedures and demographic features of patients to perform OUD prediction. However, there could be other factors such as socio-economic variables that can increase the predictive power of AI models and may be included in future models.

## Data Availability

The dataset is available from IBM at https://www.ibm.com/products/marketscan-research-databases.

## Notes

### Competing Interest Statement

The authors have declared no competing interest.

### Author Declarations

Use of this data for this study was approved as an exempt protocol by The University of Kentucky's Institutional Review Board.

## References

1. Gostin LO, Hodge JG, Noe SA. Reframing the Opioid Epidemic as a National Emergency. JAMA. 2017;318(16):1539. doi:10.1001/jama.2017.13358

2. Pitt AL, Humphreys K, Brandeau ML. Modeling Health Benefits and Harms of Public Policy Responses to the US Opioid Epidemic. Am J Public Health. 2018;108(10):1394–1400. doi:10.2105/AJPH.2018.304590

3. Substance Abuse and Mental Health Services Administration. Key Substance Use and Mental Health Indicators in the United States: Results from the 2020 National Survey on Drug Use and Health. Substance Abuse and Mental Health Services Administration; 2021. Accessed December 15, 2021. https://www.samhsa.gov/data/sites/default/files/reports/rpt35325/NSDUHFFRPDFWHTMLFiles2020/2020NSDUHFFR1PDFW102121.pdf

4. Olsen Y, Sharfstein JM. Confronting the Stigma of Opioid Use Disorder—and Its Treatment. JAMA. 2014;311(14):1393. doi:10.1001/jama.2014.2147

5. Wu LT, Zhu H, Swartz MS. Treatment utilization among persons with opioid use disorder in the United States. Drug Alcohol Depend. 2016;169:117-127.

6. Cheattle MD. Risk Assessment: Safe Opioid Prescribing Tools. Accessed December 15, 2021. https://www.practicalpainmanagement.com/resource-centers/opioid-prescribing-monitoring/risk-assessment-safe-opioid-prescribing-tools

7. Gao W, Leighton C, Chen Y, Jones J, Mistry P. Predicting opioid use disorder and associated risk factors in a Medicaid managed care population. Am J Manag Care. 2021;27(4):148–154. doi:10.37765/ajmc.2021.88617

8. Webster LR, Webster RM. Predicting Aberrant Behaviors in Opioid-Treated Patients: Preliminary Validation of the Opioid Risk Tool. Pain Med. 2005;6(6):432–442. doi:10.1111/j.1526-4637.2005.00072.x

9. Cheatle MD, Compton PA, Dhingra L, Wasser TE, O’Brien CP. Development of the Revised Opioid Risk Tool to Predict Opioid Use Disorder in Patients with Chronic Nonmalignant Pain. J Pain. 2019;20(7):842–851. doi:10.1016/j.jpain.2019.01.011

10. McNeely J, Kumar PC, Rieckmann T, et al. Barriers and facilitators affecting the implementation of substance use screening in primary care clinics: a qualitative study of patients, providers, and staff. Addict Sci Clin Pract. 13:8.

11. National Ambulatory Medical Care Survey: 2018 National Summary. Published online 2018:40.

12. Evans E, Grella CE, Murphy DA, Hser YI. Using Administrative Data for Longitudinal Substance Abuse Research. J Behav Health Serv Res. 2010;37(2):252–271. doi:10.1007/s11414-008-9125-3

13. Breiman L. Random Forests. Mach Learn. 2001;45:5-32.

14. Natekin A, Knoll A. Gradient boosting machines, a tutorial. Front Neurorobotics. 2013;7. doi:10.3389/fnbot.2013.00021

15. Tolles J, Meurer WJ. Logistic Regression: Relating Patient Characteristics to Outcomes. JAMA. 2016;316(5):533. doi:10.1001/jama.2016.7653

16. Lo-Ciganic WH, Huang JL, Zhang HH, et al. Evaluation of Machine-Learning Algorithms for Predicting Opioid Overdose Risk Among Medicare Beneficiaries With Opioid Prescriptions. JAMA Netw Open. 2019;2(3):e190968. doi:10.1001/jamanetworkopen.2019.0968

17. Lo-Ciganic WH, Huang JL, Zhang HH, et al. Using machine learning to predict risk of incident opioid use disorder among fee-for-service Medicare beneficiaries: A prognostic study. Lu K, ed. PLOS ONE. 2020;15(7):e0235981. doi:10.1371/journal.pone.0235981

18. Cochran BN, Flentje A, Heck NC, et al. Factors predicting development of opioid use disorders among individuals who receive an initial opioid prescription: Mathematical modeling using a database of commercially-insured individuals. Drug Alcohol Depend. 2014;138:202-208. doi:10.1016/j.drugalcdep.2014.02.701

19. Reps JM, Cepeda MS, Ryan PB. Wisdom of the CROUD: Development and validation of a patient-level prediction model for opioid use disorder using population-level claims data. Gullo MJ, ed. PLOS ONE. 2020;15(2):e0228632. doi:10.1371/journal.pone.0228632

20. Hasan MM, Young GJ, Patel MR, Modestino AS, Sanchez LD, Noor-E-Alam Md. A machine learning framework to predict the risk of opioid use disorder. Mach Learn Appl. 2021;6:100144. doi:10.1016/j.mlwa.2021.100144

21. Ellis RJ, Wang Z, Genes N, Ma’ayan A. Predicting opioid dependence from electronic health records with machine learning. BioData Min. 2019;12(1):3. doi:10.1186/s13040-019-0193-0

22. Wadekar AS. Understanding Opioid Use Disorder (OUD) using tree-based classifiers. Drug Alcohol Depend. 2020;208:107839. doi:10.1016/j.drugalcdep.2020.107839

23. Karhade AV, Cha TD, Fogel HA, et al. Predicting prolonged opioid prescriptions in opioid-naïve lumbar spine surgery patients. Spine J. 2020;20(6):888–895. doi:10.1016/j.spinee.2019.12.019

24. Grazal CF, Anderson AB, Booth GJ, Geiger PG, Forsberg JA, Balazs GC. A Machine-Learning Algorithm to Predict the Likelihood of Prolonged Opioid Use Following Arthroscopic Hip Surgery. Arthrosc J Arthrosc Relat Surg. Published online August 2021:S0749806321007428. doi:10.1016/j.arthro.2021.08.009

25. Ripperger M, Lotspeich SC, Wilimitis D, et al. Ensemble learning to predict opioid-related overdose using statewide prescription drug monitoring program and hospital discharge data in the state of Tennessee. J Am Med Inform Assoc. Published online October 19, 2021:ocab218. doi:10.1093/jamia/ocab218

26. Ferris LM, Saloner B, Krawczyk N, et al. Predicting Opioid Overdose Deaths Using Prescription Drug Monitoring Program Data. Am J Prev Med. 2019;57(6):e211–e217. doi:10.1016/j.amepre.2019.07.026

27. Fouladvand S, Hankosky ER, Bush H, et al. Predicting substance use disorder using long-term attention deficit hyperactivity disorder medication records in Truven. Health Informatics J. 2020;26(2):787–802. doi:10.1177/1460458219844075

28. Che Z, St. Sauver J, Liu H, Liu Y. Deep Learning Solutions for Classifying Patients on Opioid Use. In: AMIA Annual Symposium.; 2017:10.

29. Fouladvand S, Talbert J, Dwoskin LP, et al. Predicting Opioid Use Disorder from Longitudinal Healthcare Data using Multi-stream Transformer. AMIA Annu Symp. Published online November 1, 2021. Accessed December 15, 2021. http://arxiv.org/abs/2103.08800

30. Dong X, Deng J, Rashidian S, et al. Identifying risk of opioid use disorder for patients taking opioid medications with deep learning. J Am Med Inform Assoc. 2021;28(8):1683–1693. doi:10.1093/jamia/ocab043

31. Kashyap A, Callison-Burch C, Boland MR. A Deep Learning Method to Detect Opioid Prescription and Opioid Use Disorder from Electronic Health Records. Accessed December 23, 2021. https://www.medrxiv.org/content/10.1101/2021.09.13.21263524v1

32. Workman TE, Shao Y, Kupersmith J, et al. Explainable Deep Learning Applied to Understanding Opioid Use Disorder and Its Risk Factors. In: 2019 IEEE International Conference on Big Data. IEEE; 2019.

33. IBM MarketScan Research Database. Accessed December 20, 2021. https://www.ibm.com/products/marketscan-research-databases

34. Medi-Span. Accessed February 6, 2022. https://www.wolterskluwer.com/en

35. HCUP User Support (HCUP-US). HCUP User Support (HCUP-US). Accessed February 6, 2022. https://www.hcup-us.ahrq.gov/toolssoftware/ccs/ccs.jsp

36. Zaremba W, Sutskever I, Vinyals O. Recurrent Neural Network Regularization. ArXiv14092329 Cs. Published online February 19, 2015. Accessed December 19, 2021. http://arxiv.org/abs/1409.2329

37. Vaswani A, Shazeer N, Parmar N, et al. Attention is All you Need. In: Advances in Neural Information Processing Systems. Vol 30; 2017:11.

38. Lipton ZC, Kale DC, Elkan C, Wetzel R. Learning to Diagnose with LSTM Recurrent Neural Networks. ArXiv151103677 Cs. Published online March 21, 2017. Accessed February 6, 2022. http://arxiv.org/abs/1511.03677

39. Goodfellow I, Bengio Y, Courville C. Softmax Units for Multinoulli Output Distributions. In: Deep Learning. MIT; 2016:180–184.

40. Alpaydin E. Introduction to Machine Learning. MIT press; 2020.

41. Opioid Oral Morphine Milligram Equivalent (MME) Conversion Factors. Accessed December 21, 2021. https://medicaid.utah.gov/Documents/files/Opioid-Morphine-EQ-Conversion-Factors.pdf

42. Dowell D, Haegerich TM, Chou R. CDC Guideline for Prescribing Opioids for Chronic Pain—United States, 2016. JAMA. 2016;315(15):1624. doi:10.1001/jama.2016.1464

43. Dunn KM, Saunders KW, Rutter CM, et al. Overdose and prescribed opioids: Associations among chronic non-cancer pain patients. Published online 2010:16.

44. Sullivan MD, Edlund MJ, Fan MY, DeVries A, Braden JB, Martin BC. Risks for possible and probable opioid misuse among recipients of chronic opioid therapy in commercial and medicaid insurance plans: The TROUP Study. Pain. 2010;150(2):332–339. doi:10.1016/j.pain.2010.05.020

45. Edlund MJ, Martin BC, Russo JE, DeVries A, Braden JB, Sullivan MD. The Role of Opioid Prescription in Incident Opioid Abuse and Dependence Among Individuals With Chronic Noncancer Pain: The Role of Opioid Prescription. Clin J Pain. 2014;30(7):557–564. doi:10.1097/AJP.0000000000000021

46. Hser YI, Saxon AJ, Mooney LJ, et al. Escalating Opioid Dose Is Associated With Mortality: A Comparison of Patients With and Without Opioid Use Disorder. J Addict Med. 2019;13(1):41–46. doi:10.1097/ADM.0000000000000458

47. Lundberg SM, Lee SI. A Unified Approach to Interpreting Model Predictions. In: Proceedings of the 31st International Conference on Neural Information Processing Systems.; 2017:10.

48. Mark TL, Dilonardo J, Vandivort R, Miller K. Psychiatric and medical comorbidities, associated pain, and health care utilization of patients prescribed buprenorphine. J Subst Abuse Treat. 2013;44:481-487.

49. Scherrer JF, Salas J, Sullivan MD, et al. Impact of adherence to antidepressants on long-term prescription opioid use cessation. Br J Psychiatry. 18AD;12:103–111.

50. Webster L. Risk Factors for Opioid-Use Disorder and Overdose. Anesth Analg. 2017;125(5):1741–1748.

