## Supplemental Material for "A Comparative Effectiveness Study on Opioid Use Disorder Prediction Using Artificial Intelligence and Existing Risk Models"

[1]Institute for Biomedical Informatics; [2]Department of Computer Science; [3]Department of Internal Medicine; [4]Department of Pharmaceutical Sciences; [5]Department of Biostatistics; [6]Department of Psychiatry; [7]Department of Family and Community Medicine, University of Kentucky, Lexington, KY, USA; [8]American Board of Family Medicine, Lexington, KY, USA; [9]Department of Population Health Sciences, Weill Cornell Medicine, Cornell University, New York, NY, USA; [10]Stanford Center for Biomedical Informatics Research, Stanford University, Stanford, CA, USA.

### S1. Data format

For each of the enrollees in the case and control cohorts, their medications, diagnoses, procedure and demographic records between 2009 to 2020 were extracted. The original format in IBM MarketScan data is a table where each row is a visit and columns are enrollee ID, date of visit, and prescription/diagnosis/procedure details. Each data stream was converted into an enrollee-time matrix including tuples $(P, T, X)$ where $P$ is the enrollees set in our cohorts, $T$ is a set of monthly time points from January 2009 to June 2020 (138 time steps in total), and $X$ is the feature set for each data stream. For medications stream, $X$ includes 50 high level medication codes. For diagnoses, $X$ is a set of 138 diagnoses CCS codes. Procedures includes 79 procedures CCS codes. This dynamic data was used to train the LSTM, transformer and MUPOD. Further, we created a static data to train classical machine learning models including logistic regression, random forest and XGBoost as these models are more effective when trained using static data rather than sparse and high dimensional temporal data. Therefore, longitudinal data $X(P, T, X)$ were converted to a new format including tuples $(P, F)$, where $P$ is the complete list of enrollees, and $F$ includes frequency values for all medication, diagnosis, and procedures across the time steps concatenated with demographic features age and sex.

### S2. Anchor based cohort matching

Cases and controls were matched based on age, sex, monthly opioid prescription use and longitudinal data availability using an anchor-based method. For each sex group in the case cohort, patients were first clustered into multiple groups using k-means clustering algorithm. Age, sex, monthly opioid prescription use and longitudinal data availability were used for clustering the cases. The number of clusters were tuned using elbow method[1] (10 cluster for males and 11 clusters for females; Figure S1). These cluster centers combined with one percent of randomly selected samples from each cluster formed a set of 1,032 and 1,360 anchors for male and female cohorts, respectively. Cases and controls were then matched based on their average distance to these anchors. In fact, for each case $x$ with average distance of $d$ from the corresponding (males or female) anchors, a control sample $y$ with an average distance of $d \pm e$ to the anchors was selected where $e$ is the minimum value across all non-matched control samples.

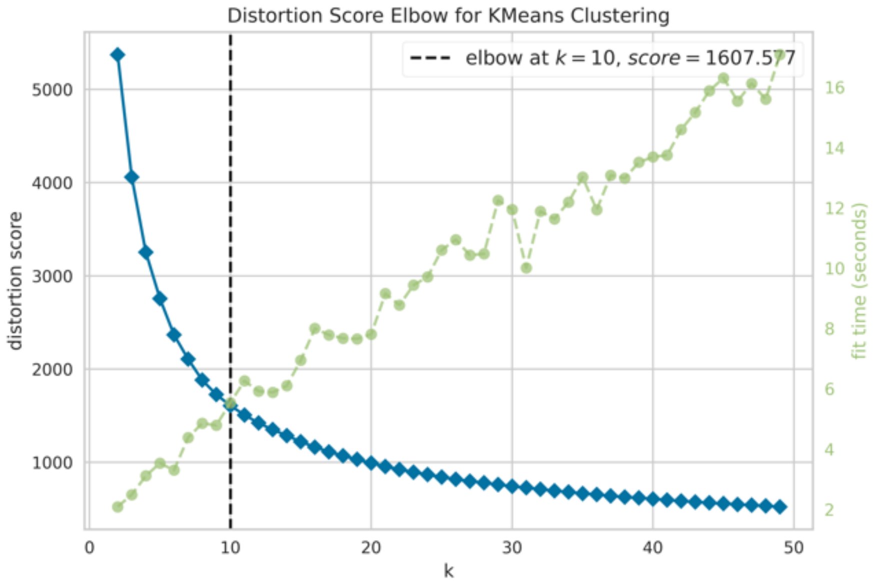


(a) Male patients

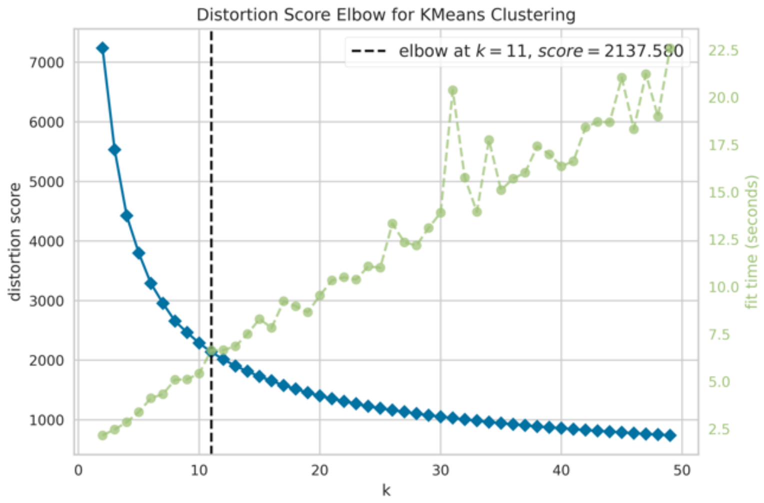


(b) Female patients

Figure S1. Distortion score vs number of clusters for both female and male cases. The dashed black line shows the optimum number of clusters for each cohort.

Figure S2 shows the effect of this matching on case and control cohorts. Note, this figure is created using 2K randomly selected samples for clarity and visualization purpose. In Figure S2.a, the longitudinal medications, diagnoses and procedures data were first converted to a static data. Dimensionality of this static data was then reduced to 2D using tSNE algorithm[2]. This figure shows that cases (red dots) and controls (blue dots) distributions are well matched in the feature space. Together cases and controls can train a machine learning model efficiently to discriminate between cases and controls. Figure S2.b shows the distributions of opioid use frequencies for cases (red bars) and controls (blue bars). X-axis in this figure shows the frequency of opioid use and y-axis is the number of patients with the relevant opioid use frequency. As opioid use increases (toward the tail of the histogram), the number of cases (red bar) increases in comparison to controls (Figure S2.b).

However, cases and controls are still well matched as the distributions are similar across x-axis.

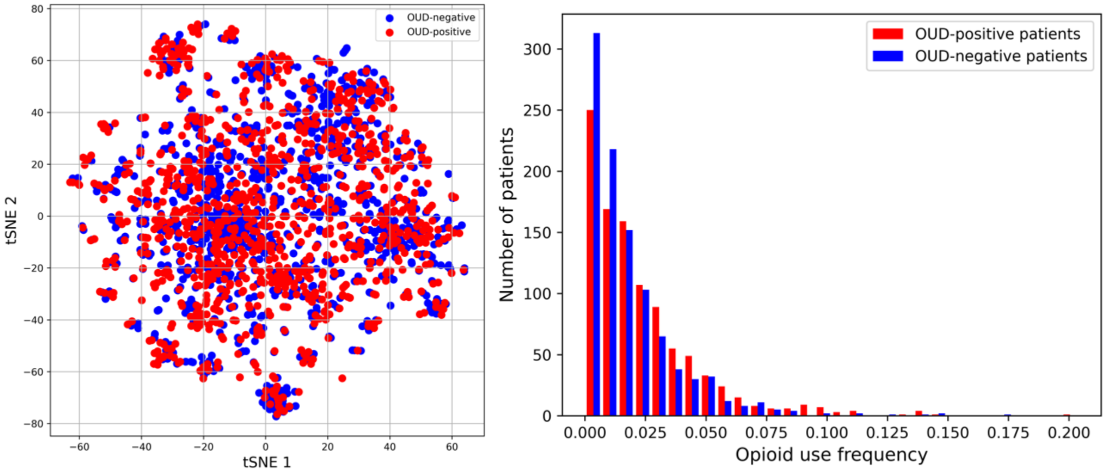


(a) Overall distribution. (b) Opioid use duration.

Figure S2. Effect of case and control matching.

### S3. Feature filtering

Original feature set includes 619 predictors (94 medications, 283 diagnoses and 242 procedure high level codes). However, many of these features are rare events causing a high level of sparsity in the data, which can significantly reduce the efficacy of the AI models[3]. Figure S3.a, b and c show the sparsity distributions for medications, diagnoses and procedures, respectively. X-axis shows features (each bar is associated with one feature) and y-axis shows the number of patients for whom that predictor value is non-zero at least during one visit. Assuming the distributions are half-bell, we excluded features that were more than two standard deviation from mean of the full bell shape distribution. As a result, the final predictor set includes 269 variables (50 medications, 138 diagnoses, 79 procedures plus 2 demographic features).

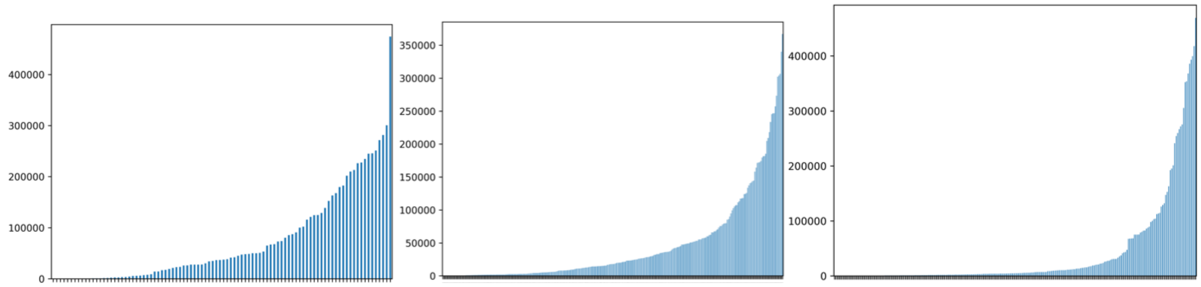


(a) Medications (b) Diagnoses (c) Procedures

Figure S3. Sparsity distribution of medications, diagnoses and procedures variables.

### S5. Multi-stream transformer model architecture

#### *Data representation*

Multi-stream transformer for opioid use disorder prediction (MUPOD) is a transformer-based deep learning model that we have designed to analyze $n$ highly correlated healthcare data streams simultaneously. To minimize ambiguity, the algorithm is described for a single patient and for $n = 3$. Each patient can be represented by $p = (S, y)$ in which $S$ is a set of input streams and $y$ is the target variable. Herein, three input streams are considered: 1) medication tuples $(T, M)$ in which $t_i$ is the $i^{th}$ time step and $M$ is a list of 50 medication codes represented as a multi-hot encoding vector at time $t_i$, 2), diagnoses tuples $(T, D)$ where $t_i$ is the $i^{th}$ time step and $D$ is a list of 138 diagnoses CCS codes represented as a multi-hot encoding vector for patient $P$ at time $t_i$, 3) procedures tuples $(T, P)$ in which $t_i$ is the $i^{th}$ time step and $P$ is a set of 79 procedure codes represented as a multi-hot encoding vector for the patient $P$ at $t_i$. These data streams, $(T, M)$ and $(T, D)$ and $(T, P)$, are represented using LSTM models to address sparsity and high dimensionality in the raw data.

The goal of data representation is to learn a function: $f_R: X \rightarrow \mathbb{R}^d$ where $d$ shows the dimension of the representation to which each input stream is mapped, $X \in \{M, D, P\}$, and $M$, $D$ and $P$ are medication, diagnosis, and procedure streams, respectively. To train $f_R$, three LSTM were separately trained using medications $(T, M)$, diagnosis $(T, D)$ and procedure $(T, P)$ streams. The intermediate outputs from the trained LSTM hidden states were used to represent case and control cohorts. The general schema of the data pre-processing and representation is shown in Figure S4.

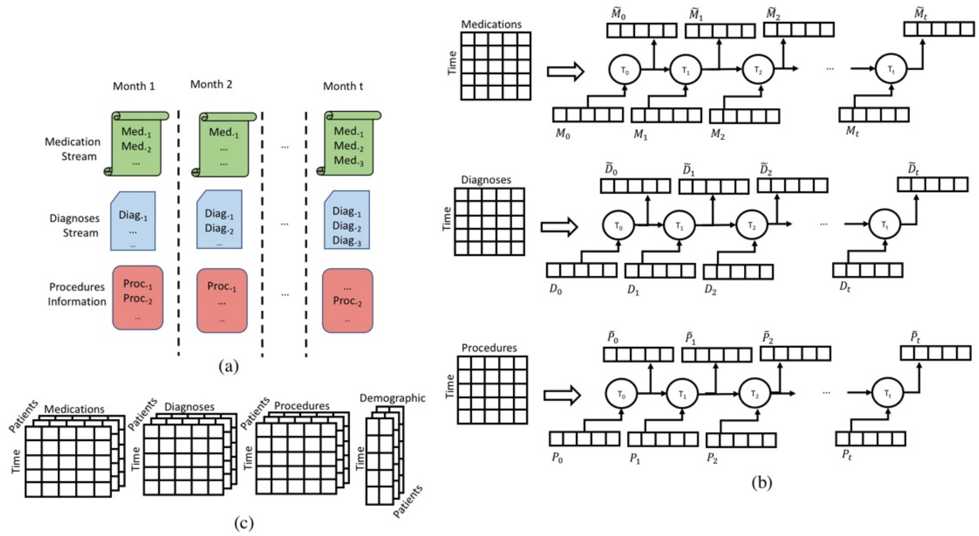


Figure S4. Data representation. Data is first converted to an enrollee-time matrix $X(P, T, F)$. Then, each stream of data is fed to a different LSTM model to produce the represented data.

***Multi-stream self-attention layer***

The represented input streams, $M$, $D$, and $P$, are first fed into the self-attention layer to generate query, key, and value matrices for each input stream. For example, medications $M$ are fed to a set of fully connected layers to generate $M_Q$, $M_K$, and $M_V$, representing the query, key, and value matrices for the represented medication stream for patient $P$. Let $X, Y \in \{M, D, P\}$, the query, key, and value matrices are used to find the attentions across these three input streams:

$$Attention(X_Q, Y_K, Y_V) = softmax(\frac{X_Q Y_K^T}{\sqrt{d_k}})Y_V \quad (1)$$

Note, the $d_k$ is the same as the original transformer. Figure S5 shows the general architecture of MUPOD and a detailed description of MUPOD's self-attention layer. The raw medication and diagnose streams are first represented in the representation layer (the intermediate outputs of the LSTMS models in Figure S4). The temporal information is then encoded into the represented streams using the original position encoding in transformer. The encoded streams are processed in the MUPOD's multi-stream encoder layer. This novel multi-attention layer is further described in more details in Figure 5.b. In the figure, $X_Q$, $X_K$, and $X_V$ represent query, key, and value matrices for stream $X (X \in \{M, D, P\})$. All possible combinations of the data streams are used to determine the attention weights between different time points and across streams. Attentions are then passed through a set of dense layers to generate outputs. Given three data streams $M$, $D$ and $P$, six combinations can be generated i.e., $MM$, $MD$, $MP$, $DD$, $DP$ and $PP$.

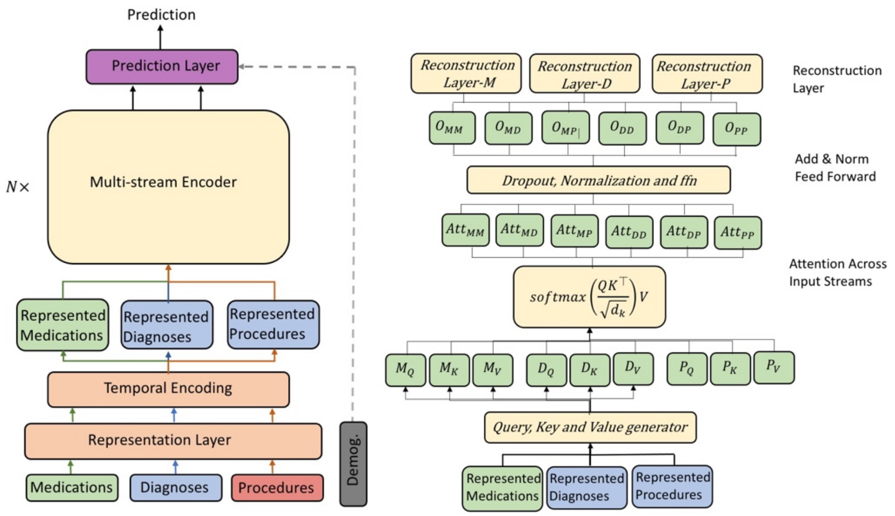


(a) General architecture of MUPOD (b) Multis-stream self

Figure S5. MUPOD architecture. $X_Q$, $X_K$, and $X_V$ represent query, key, and value matrices for the input stream $X$, where $X \in \{M, D, P\}$. $Att_{XY}$ represents the attention weights between different records across input streams $X$ and $Y$, where $X, Y \in \{M, D, P\}$. $O_{XY}$ represents the outputs, which capture the associations between the input streams $X$ and $Y$. The demographic information is plugged into the system before the last layer and in the classification layer.

The reconstruction layer receives the relevant outputs and maps them to appropriate format for the next layer as described in Equation 2. For example, only the outputs relevant to the medications ($M$) including $MM$, $MD$ and $MP$ are used to reconstruct the medication stream:

$$f: O_{XX}, O_{XY} \rightarrow \hat{X}$$

$$\hat{X} = [concat(O_{XX}, O_{XY})]W_x + b_x \quad (2)$$

where $O_{XX}, O_{XY} \subset \{O_{MM}, O_{MD}, O_{DP}, O_{DD}, O_{PP}\}$, $\hat{X} \in \{\widehat{M}, \widehat{D}, \hat{P}\}$, $X, Y \in \{M, D, P\}$, $W_x$ and $b_x$ are trainable reconstruction weight and bias matrices. The two reconstructed matrices generated by the last encoder layer are fed to classification layer to make the final decision for the current patient $p$ as $Softmax([concat(\widehat{M}, \widehat{D}, \hat{P})]W + b)$.

### S6. Settings

The data set include 474,208 patients, which was randomly grouped into 426,812 training samples and 100 randomly selected testing sets each with 1% of the test cohort of 47,396 patients in total. Note, none of the test sets overlap with the train set. Logistic regression, random forest and XGboost models were trained using the train set and optimized using a randomized search 3-fold cross validation. The optimum logistic regression has a sag as its solver with no penalty and $C = 0.0001$. The optimum random forest model includes 1800 estimators with maximum depth of 60. The optimum values for the number of estimators, maximum depth, learning rate and gamma for the XGBoost model used in Tables 2 and 3 are 2000, 64, 0.01, and 0.1, respectively. Note, for the risk identification results presented in Figure 1, we used a smaller XGBoost model due to high complexity of the creating SHAP trees using our big data. This smaller XGBoost model was trained using randomly selected five percent of the data (23,711 samples) and top-10 frequently used medications, diagnoses and procedures variables plus the demographic information including age and sex. The accuracy, precision, recall, F1-score and AUC for this smaller XGBoost model are 0.598, 0.595, 0.611, 0.603 and 0.643, respectively. Deep learning models were trained using pre-defined hyper-parameters due to the high time complexity of the training process. The learning rate, regularization factor, number of hidden neurons, number of epochs and batch size for the LSTM model are equal to 0.01, $10^{-6}$, 48, 1 and 256, respectively. The transformer model includes 4 layers trained with a learning rate, dropout rate, number of epochs, number of heads, and batch size of 0.001, 0.3, 5, 4 and 256, respectively. The number of layers, learning rate, dropout rate, number of epochs, number of heads and batch size for MUPOD were 4, 0.001, 0.3, 5, 4 and 256, respectively. All models were trained on a server machine equipped with eight GeForce GTX 1080 GPUs.
